# Explainable, federated deep learning model predicts disease progression risk of cutaneous squamous cell carcinoma

**DOI:** 10.1101/2024.08.22.24312403

**Authors:** Juan I. Pisula, Doris Helbig, Lucas Sancéré, Oana-Diana Persa, Corinna Bürger, Anne Fröhlich, Carina Lorenz, Sandra Bingmann, Dennis Niebel, Konstantin Drexler, Jennifer Landsberg, Roman Thomas, Katarzyna Bozek, Johannes Brägelmann

**Author notes:** Equal contribution (Juan I. Pisula, Doris Helbig, Katarzyna Bozek, Johannes Brägelmann).

## Abstract

Predicting cancer patient disease progression is a key step towards personalized medicine and secondary prevention. The ability to predict which patients are at an elevated risk of developing local recurrences or metastases would allow for tailored surveillance of these high-risk patients as well as enhanced and timely interventions.

We developed a deep learning transformer-based approach for prediction of progression of cutaneous squamous cell carcinoma (cSCC) patients based on diagnostic histopathology slides of the tumor. Our model, trained in a federated manner on patient cohorts from three clinical centers, reached an accuracy of AUROC=0.82, surpassing the predictive power of clinico-pathological parameters used to assess progression risk. We conducted an interpretability analysis, systematically comparing a broad range of spatial and morphological features that characterize tissue regions predictive of patient progression. Our findings suggest that information located at the tumor boundaries is predictive of patient progression and that heterogeneity of tissue morphology and organization are characteristic of progressive cSCCs. Trained in a federated fashion exclusively on standard diagnostic slides obtained during routine care of cSCC patients, our model can be deployed and expanded across other clinical centers. This approach thereby offers a potentially powerful tool for improved screening and thus better clinical management of cSCC patients.

## Introduction

Cutaneous squamous cell carcinoma (cSCC) is the second most prevalent type of non-melanoma skin cancer that is diagnosed in 1 million patients in the USA every year.^1^ In the last decades, the incidence of cSCC has risen sharply and is projected to increase further.^2^ Even though the majority of cSCCs can be removed by surgical excision, a relevant fraction of patients experience disease progression by local recurrence or metastases to lymph nodes or other body sites, which is associated with poor prognosis and increased risk of death.^3–6^ Due to the high incidence of cSCC, this poses a significant public health concern. Reliable predictors are thus needed to decide which patients will benefit from enhanced secondary prevention e.g. by more frequent follow-up care or additional treatments such as immuno-, chemo- or radiotherapy. Current cSCC staging systems like the American Joint Committee on Cancer (AJCC), the Brigham Women’s Hospital (BWH), or the National Comprehensive Cancer Network (NCCN) staging systems aim to provide guidance on risk stratification and clinical management of cSCC patients.^7,8^ However, they fall short of reliably identifying patients at high risk of disease progression. Recently, multi-gene expression signatures have been used to predict metastasis risk of cSCCs.^9,10^ While these signatures help to predict metastasis risk, they have not yet been used to predict local recurrences. In addition, they require measurement of gene expression from patient samples, which limits their potential for translation into clinical routine use.

In addition to clinical parameters such as immunosuppression, several pathological tumor features such as perineural involvement, tumor size, and invasion depth have been associated with increased risk of cSCC progression.^4–6^ Moreover, specific histological subtypes e.g. desmoplastic cSCC have been linked to higher recurrence and/or metastasis risk.^6^ Morphology in histological specimens thus holds information on progression risk, but has not yet been exploited systematically. Since deep learning has matched human experts in cancer detection and classification,^11^ computational pathology methods hold promise to extract information on patient progression from histopathology image data. Building robust models that offer high predictive power across data independent of their source, requires multi-institutional data sets for model training. Obtaining such data sets poses challenges regarding data governance and raises concerns about patient privacy. Federated Learning (FL) is a strategy that limits the logistic overhead and reduces privacy concerns in training a multi-center-based model.^12,13^ Moreover, FL simplifies the inclusion of new patients and cohorts for further model training, which in turn facilitates model update, continuous improvement, and clinical applicability.

Here, we present a multiple instance learning transformer-based deep learning model for prediction cSCC progression risk using Hematoxylin-Eosin-(HE-) stained histopathology images acquired during routine care (**Fig. 1**).^14,15^ Our model, trained in a federated manner on cohorts from three clinical centers, achieved high accuracy in predicting patients at risk of disease progression, which corresponds to significant differences in progression-free survival. We developed explainability methods on our model which provide insights into the tissue areas and cell features associated with increased progression risk. Overall, we present a powerful approach that improves risk-stratification of cSCC patients and offers insights into the underlying cancer biology.

**Figure 1:**
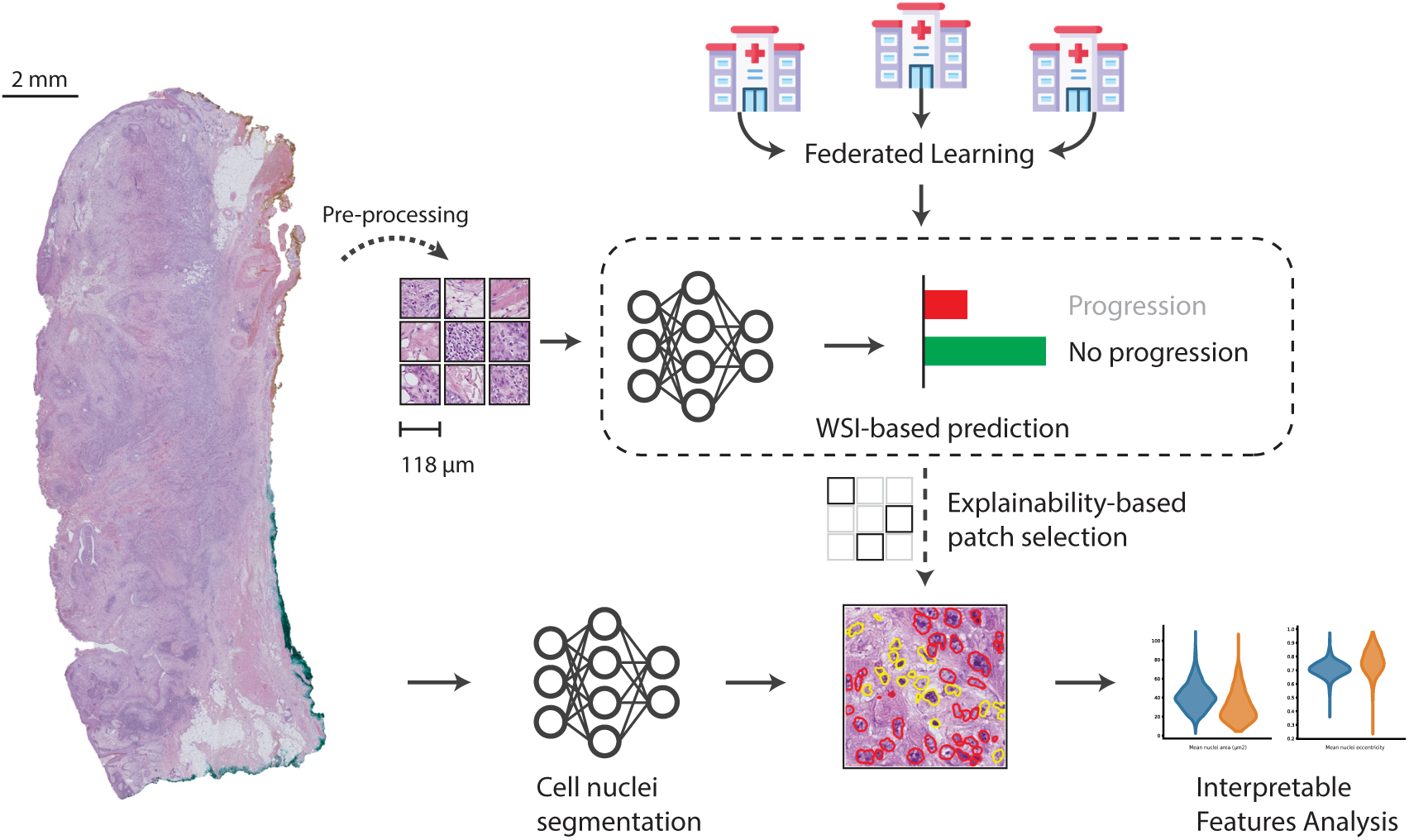
We propose a WSI-based cutaneous Squamous Cell Carcinoma (cSCC) progression prediction model, trained on data from three medical centers using Federated Learning. Beyond prediction, we investigate underlying biological features that influence our classifier. We do so by computing cellular-level features with aid of a nuclei segmentation model. We analyze these features in image regions detected as relevant for prediction outcome by Integrated Gradients, an input attribution algorithm for explainable deep neural networks.

## Results

### Deep learning on histopathology images predicts cSCC progression risk

Currently, it is not clear if the progression risk of a cSCC can be inferred from a histopathology slide and if so, which elements of the tumor and its microenvironment are decisive of disease progression. To fill this gap, we used a multiple instance learning, transformer-based classifier for the task of progression prediction from Whole Slide Images (WSIs). We trained the model in a federated manner, leveraging data from three different medical centers (**Fig. 1**).^14,15^

Initially, we trained our model on the Cologne cohort only (n=157 patients, 214 WSIs), achieving cSCC progression status classification accuracy of 0.92 AUROC (95% CI=[0.83-1.00]) in a held-out test set from Cologne (**Fig. 2A)**. In comparison, a multivariable logistic regression model incorporating clinico-pathological parameters associated with risk of disease progression (**Suppl. Fig. 1**) achieved an AUROC of 0.64 (95% CI=[0.52-0.75]) in the same prediction task and cohort (**Fig. 2B**). To test the robustness of our deep learning model we assembled two additional cohorts from dermatology departments at the University Hospital Bonn (Bonn cohort, n=35 patients, 133 WSIs) and the Technical University Munich (Munich cohort, n=51 patients, 113 WSIs). While the model trained on the Cologne cohort performed well on the Bonn cohort (AUROC=0.90, 95% CI=[0.71-0.97]), it failed to generalize to the Munich cohort (AUROC=0.46, 95% CI=[0.30-0.63]; **Fig. 2A**). This highlights that variation induced by e.g. technical procedures or distribution shift and domain adaptation problems may hamper generalizability of models trained on a single-center cohort.

**Figure 2:**
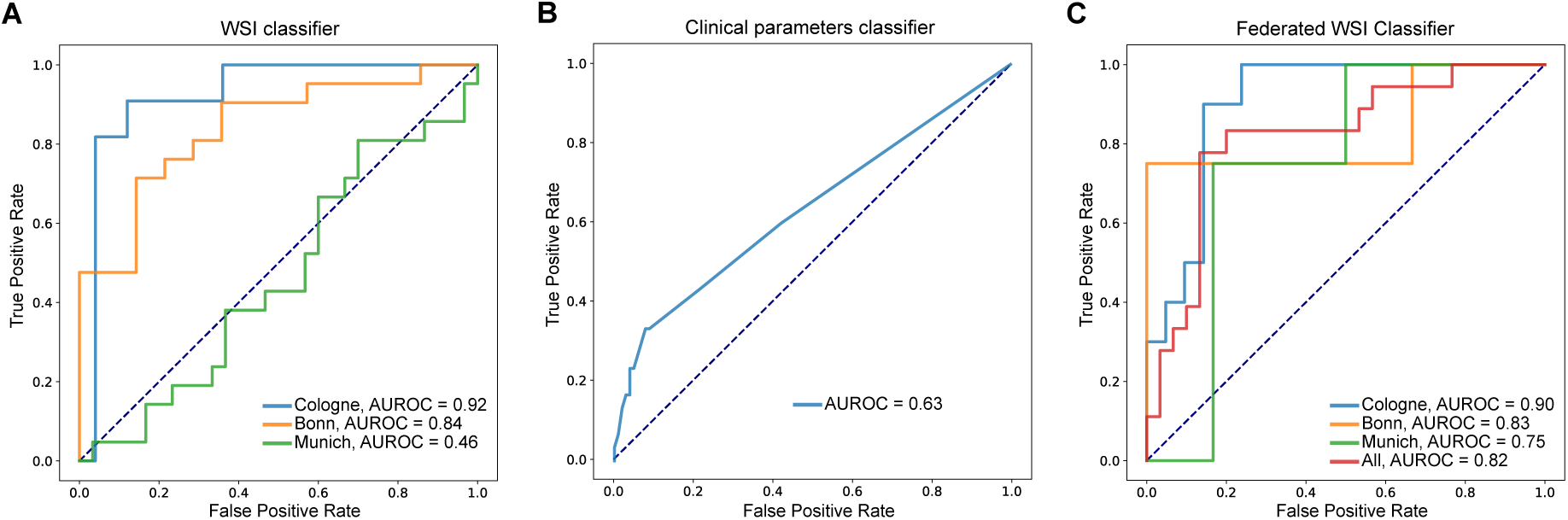
ROC curves of the classifiers. **A**: WSI-based classifier trained exclusively on the Cologne cohort and tested on Munich and Bonn cohorts (AUROC = Area under the receiver operator curve). **B**: Multivariate logistic regression model based on clinico-pathological parameters associated with progression risk in univariate analysis. Model trained and evaluated on the Cologne cohort. **C**: Federated WSI-based classifier.

### Federated learning improves generalizability of image-based classification

To improve performance across cohorts, it is crucial to train deep learning models on large and diverse datasets. However transfer of patient data and histological slides across hospitals carries important logistic complexity and poses potential privacy threats. We therefore trained our model in an FL scheme on all three cohorts (**Fig. 1**).^12^ FL overcomes the data sharing hurdles by reducing the organizational overhead of combining different patient cohorts, since patient data can remain in the respective hospital. Model training is performed locally and only model parameters are shared between the hospitals. Moreover, it enables dynamic patient enrollment and facilitates inclusion of additional centers, which in turn increases its flexibility and the opportunities for clinical deployment. Training on the multi-institutional cohorts using the FL framework did indeed improve model performance. While AUROC on Cologne and Bonn decreased at most by 2%, performance on the Munich cohort increased by 63%, leading to prediction accuracy of AUROC=0.82 (95% CI=[0.69-0.95]) in the complete dataset (**Fig. 2C**). This highlights that prediction of disease trajectories is indeed possible for cSCC patients and can be achieved with a deep learning model trained on different cohorts in a federated manner. Such prediction opens possibilities for clinical translation of the model as a tool for the identification of patients at high recurrence risk that may benefit from increased surveillance.

### Explainability analyses highlight factors associated with cSCC progression

In addition to stratifying patients according to their disease progression risk, we assessed which parts of the histological images are predictive of disease progression. We used Integrated Gradients (IGs) attributions to infer which areas in the WSIs are the most relevant for the prediction of the respective patient as progressor/non-progressor.^16^ Additionally, we leveraged a pipeline we recently established specifically for cSCC, which performs nuclei segmentation and classification of cells into one of six cell types (granulocyte, lymphocyte, plasma, stroma, tumor, and epithelial cell).^17^ We used the cell type detection and classification to analyze the WSI regions with the highest predicted power as attributed by IGs. In the WSI regions with high IGs attribution score we calculated various features of nuclei morphology, cell type composition and spatial distribution (**Suppl. Table 1**).

We next performed statistical analyses of these features to gain insights into the determinants of cSCC progression. Interestingly, many of the predictive tiles with the highest attribution score for disease progression were outside of the tumor region (**Fig. 3A**). In fact, attribution scores were low in areas with high tumor cell density, as determined using our cell type classification pipeline (**Fig. 3A, bottom left & middle**).^17^ Instead, they were high at the tumor border and frequently in areas where the most common cell type was stroma (**Fig. 3A, bottom right, Suppl. Fig. 2**).

**Figure 3:**
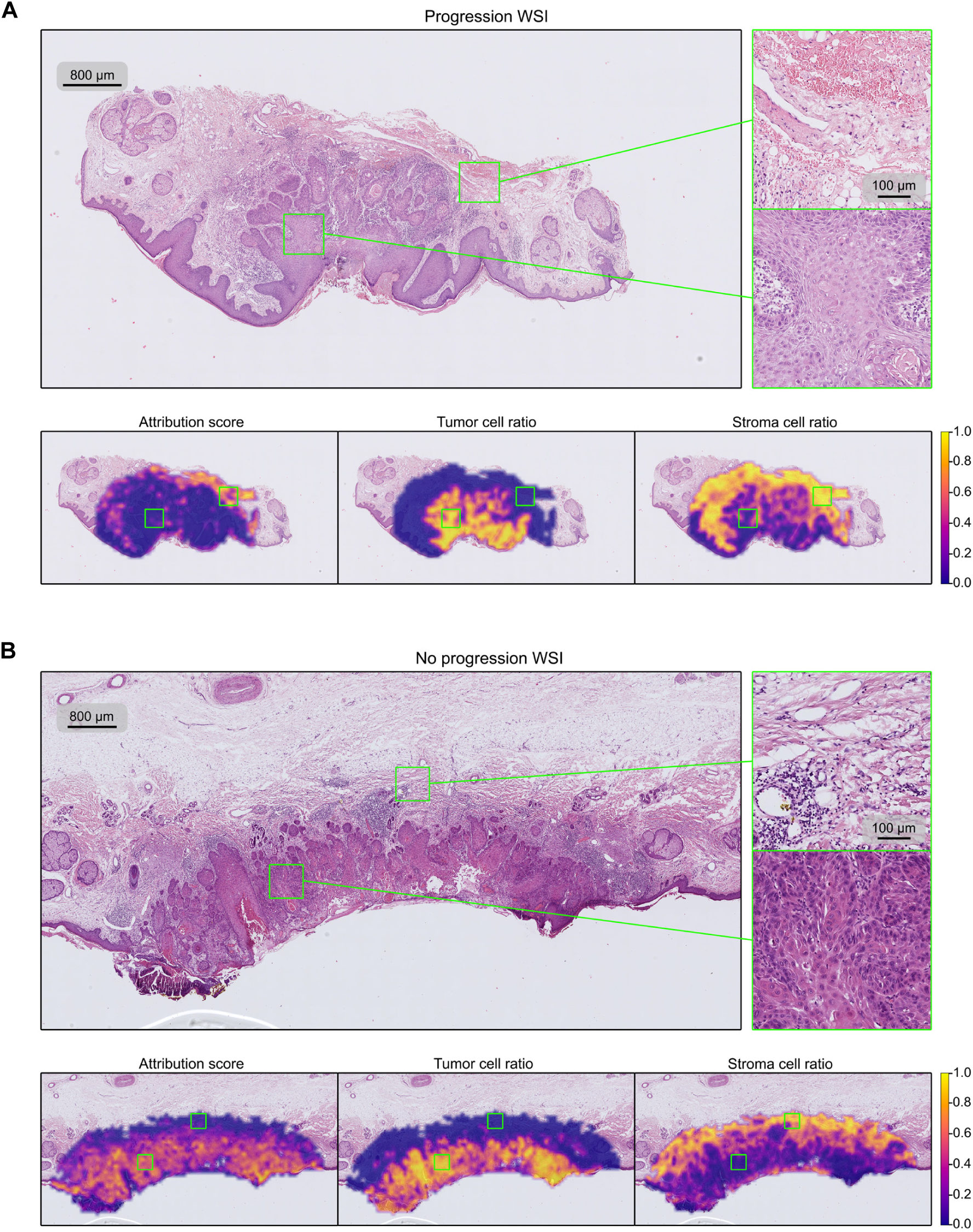
Slides and heatmaps of the patches’ classifier attribution score, tumor cell ratio, and stroma cell ratio. **A:** Slide of a progression patient, showing that the WSI-based classifier assigns higher importance to the region outside the tumor area (indicated by the tumor cell ratio heatmap). **B:** Slide of a non-progression patient, where the high attribution area coincides with the tumor-cell populated areas. Colorbar indicates the slide-normalized heatmap values.

In contrast, for patients without disease progression, the most predictive tiles were located within the tumor and in areas with high tumor cell density. Areas outside the tumor border were, in the case of these patients, not of high value for prediction of non-progression (**Fig. 3B**). This highlights that different parts of histological sections contain information that distinguishes patients at high vs. low risk of disease progression and that such patient stratification needs to be based not only on the tumor but also its surroundings for adequate predictions.

Additionally, we systematically compared the cell-based features between the tiles that were regarded as most predictive for disease progression or non-progression according to their IGs scores. Numerous parameters with significantly different distributions between the two groups were detected, **Fig. 4** shows a subset of the tumor-cell-related features. Non-progressors e.g. showed higher values in Average Nearest Neighbor Ratio (ANNR), indicating a higher uniformity in the way tumor cells were distributed (**Fig. 4A, p<0.0001**), while progressors had more intermixing of tumor cells with other cell types, i.e. more heterogeneity in tissue composition (**Fig. 4C, p<0.0001**). Moreover, tumor cells of non-progressors showed differences in their morphology compared to progressors such as larger nucleus size (**Fig. 4B, p<0.0001)** and lower nuclear eccentricity (**Fig. 4D**, **p****<0.0001**). In addition, tumors of patients that later experienced disease progression showed higher degrees of nuclear dysmorphia and pleomorphism compared to non-progressors. Tumor cells from non-progressors have larger values of morphological solidity and extent (larger median, negatively-skewed distributions, **Suppl. Fig 3A-D, Suppl. Table 1**), while morphological extent has a larger variance in tumor cells from progressors **(Suppl. Fig 3E, Suppl. Table 1)**.

**Figure 4:**
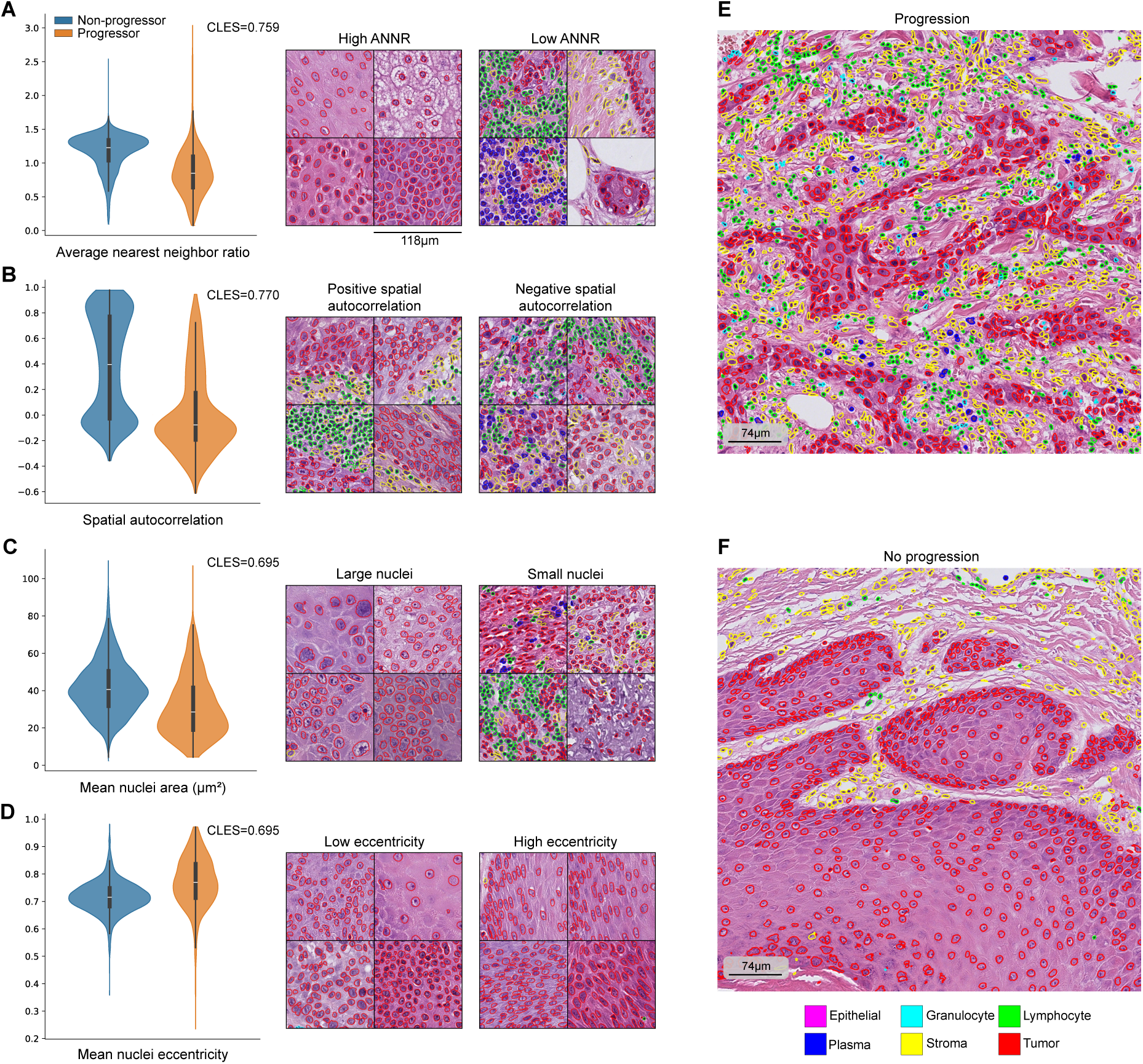
Four of the features of the tumor cells used in the analysis. **A,B,C,D** show violin plots and segmented image patches that illustrate these values. In general, progression-associated tumor cells cluster together **(A)**, interface with other cell types **(B)**, and have smaller **(C)**, eccentric nuclei **(D)**. These effects are not just local to image patches, but they occur in larger regions, as shown in **E**,**F**. The displayed CLES (Common Language Effect Size) values are indicated for the group with the largest mean. All features are significantly different in both groups, with p-values < 0.0001 using Mann-Whitney U test.

We next tested whether our cell-based features are sufficient to predict the progression/non-progression of patients based on their respective image tiles using a tree-based classification algorithm XGBoost.^18^ Interestingly, using the cell-based features as input resulted in high prediction accuracy (**Suppl. Fig. 4,** AUROC=0.98, 95% CI=[0.97-0.99]). This highlights that these features, which we computed using an independent pipeline, do indeed capture relevant biological parameters and variation associated with progression risk of patients. Thus the cellular and morphological features are making explicit the morphological and structural components of the tissues and cells that the deep learning model learned implicitly.

Overall, our explainability analyses indicate that tumor cell-intrinsic properties as well as composition of the microenvironment and growth patterns of the tumor are associated with the difference in prognosis and are captured by our deep learning model to accurately predict progression risk.

### Image-encoded information has higher discriminative power than clinical variables

Several clinico-pathological parameters have been associated with increased risk of disease progression, such as immunosuppression, perineural involvement, tumor size, and invasion depth.^4,6^ Similarly, desmoplastic cSCC histology has been linked to higher recurrence and/or metastasis risk.^6^ We used the clinico-pathological parameters available for the Cologne cohort to test their associations with survival and to compare their predictive power with the accuracy of the deep learning model. In this experiment, we used the logit output of the deep learning model as a progression risk score. Among clinico-pathological parameters, perineural invasion and beyond subcutaneous invasion were significantly associated with shorter progression free survival in univariate analyses (**Suppl. Fig. 5A, B**). Other parameters such as thickness >6mm, ulceration, and higher grade showed trends towards shorter survival, but did not reach significance (**Suppl. Fig. 5A**). Even among high-risk patients with perineural invasion or invasion beyond subcutaneous tissue not all patients developed disease progression, i.e. recurrence or metastasis. On the other hand, among the patients with one of those risk factors our deep learning model correctly separated those who progressed from those who did not based on their predicted progression risk (**Fig. 5A**). Similarly, deep learning-based predicted risk scores were higher for patients that experienced disease progression independent of tumor thickness and across histological grades (**Suppl. Fig. 5C**). The model thus allows further differentiation of patients compared to clinico-pathology-based risk factors. Without explicitly measuring these risk factors our model encodes information allowing to differentiate the two groups of patients with increased accuracy.

**Figure 5:**
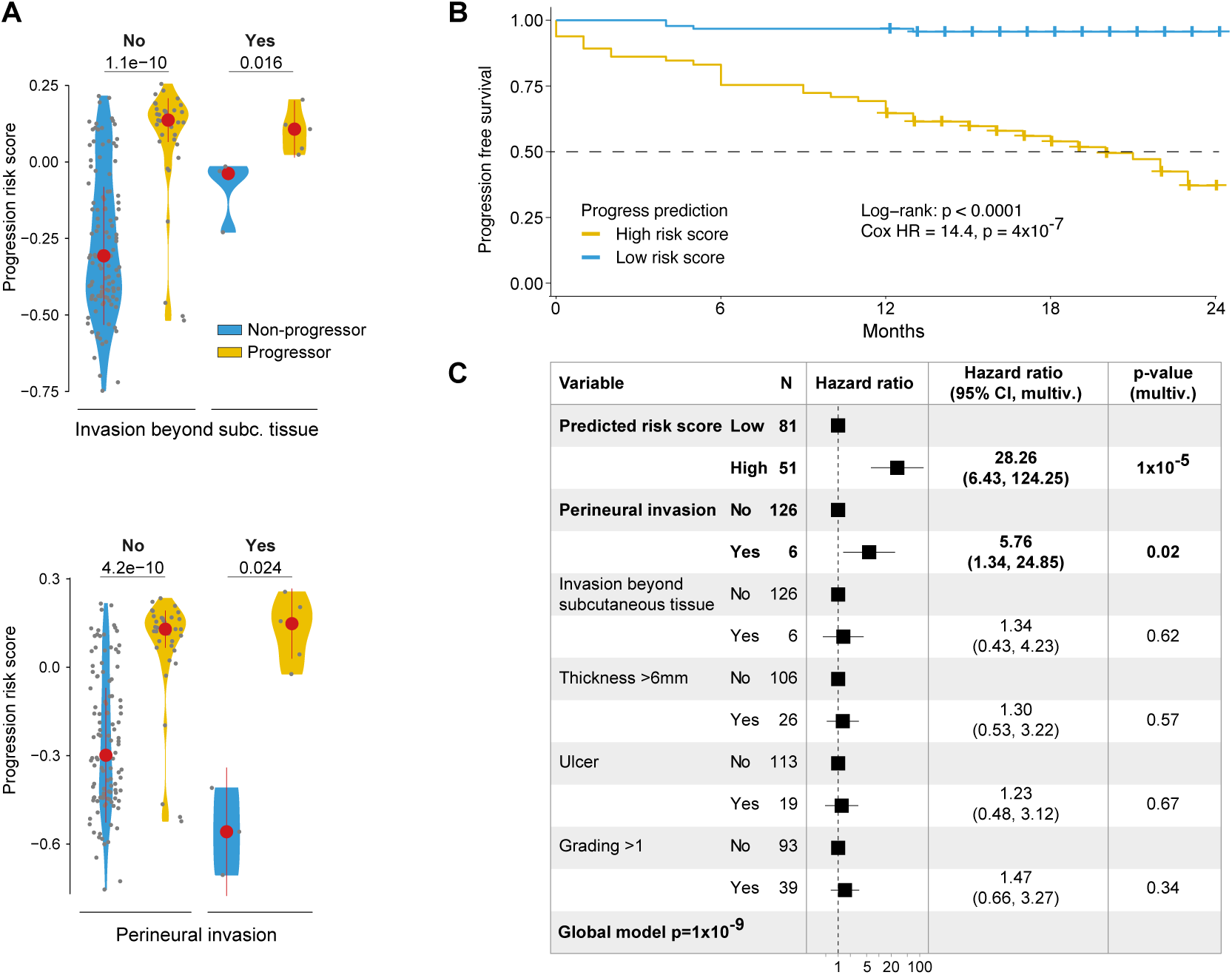
Comparison of deep learning-based classification with clinico-pathological parameters. **A:** Comparison of deep learning-based progression risk scores in Cologne patients with or without cSCC progression stratified by presence of invasion beyond subcutaneous tissue (top) or perineural invasion (bottom). Shown are median and median absolute deviation. p-values calculated by t test. **B:** Progression free survival of patients classified as high vs. low progression risk based on deep learning-based risk prediction. (Threshold determined by Youden index). Hazard ratio (HR) between groups calculated using univariate Cor regression model. **C:** Multivariable Cox regression model for n=132 Cologne patients with available data combining deep-learning based risk category with clinical parameters associated with progression free survival in univariate analyses. Shown are Hazard ratios, 95% Confidence intervals (CIs) and multivariate p-values.

We next inspected the relationship of predicted risk of progression to patient survival. Using only the deep learning model’s predicted progression risk score to classify patients as high- vs. low-risk, stratifies patients with short from those with long survival (median PFS 20.1 months vs. not reached in high vs. low risk, respectively; **Fig. 5B**). The risk of progression was 14 times higher for high- compared to low-risk patients (Hazard ratio 14.4, **p<0.0001**; **Fig. 5B**). Similarly, dividing patients into risk groups based on tertiles of the deep learning-based progression risk scores reaches similar performance (**Suppl. Fig. 5D**).

Lastly, we joined the informative factors of our clinico-pathological parameters with the deep learning model’s output to predict survival using a multivariable model. To this end we combined the deep learning model’s predicted risk scores and clinico-pathological parameters that showed a p-value below 0.1 in univariate analyses (perineural invasion, invasion beyond subcutaneous tissue, thickness >6mm, ulceration and differentiation grade >1) in a multivariable Cox regression model. This combined model showed that the image data carries more information than the clinico-pathological variables (**global p<0.0001**, **Fig. 5C**). In fact, high-risk classification based on the deep learning model carries a hazard ratio of 28.3 (**multivariable p<0.0001**). In contrast, only perineural invasion remains significant with a hazard ratio of 5.8 (multivariable p=0.02), while the other variables are non-significant (**Fig. 5C**).

Considering that only a fraction of patients is positive for perineural invasion and that clinico- pathological information is frequently incomplete, these analyses highlight the potential of our image-based model to reliably identify patients at high risk of disease progression for intensified clinical follow-up.

## Discussion

Deep learning has enabled automation of the analysis of large histopathology images. These digital pathology methods not only provide fast and detailed insights into the cellular composition of massive WSIs,^19,20^ but also allow to identify patterns and anomalies that may be imperceptible to the human eye.^21^ Here we present an approach that combines both: a model that detects complex, imperceptible morphological features of a tumor sample that are predictive of patient outcome with an explainability procedure to disentangle what these features are. While patient outcomes might be influenced by multifactorial clinical variables and span variable development trajectories, we demonstrate that, in case of cSCC, prediction of patient progression is possible based on histological images of their tumor samples alone. Via a comprehensive and quantitative analysis of predictive regions of the tumor samples we point to consistent and repetitive patterns in tumor and tumor microenvironment morphology and organization that characterize progression and non-progression patient groups. Our model offers unmatched accuracy compared to the prediction based on clinico- pathological features that were the gold standard up till now.

Our analysis combined data from three academic clinical centers: Cologne, Munich, and Bonn. The model trained on a single cohort resulted in an uneven accuracy on the remaining two cohorts, ranging from random predictions to 0.84 AUROC. While digital pathology models require large and multi-center data for better generalization, clinical data sharing carries important administrative hurdles and data protection risks. Here we demonstrate that these difficulties can be overcome by employing an FL training scheme resulting in a model with high accuracy across all cohorts while circumventing cross-center data sharing. Our model development strategy allows for easy incorporation of additional clinical centers in the future which could potentially improve the prediction accuracy further.

Deep learning models have achieved human expert-level accuracy in standard diagnostic tasks such as tumor metastases detection and cancer subtyping.^22–24^ These tasks involve detecting patterns that, while sometimes local, subtle, and difficult to notice, are known and described in pathology textbooks. In contrast, prediction of patient progression based on WSIs is a more challenging task as there are no known visual biomarkers that reliably indicate disease advancement. Numerous studies address prediction of cancer progression based on HE-stained samples of tumors across diverse tissue types,^25–30^ however rarely reaching accuracy > 0.80 AUROC. Notably, combining image with clinical data has improved prediction accuracy in some studies still barely exceeding 0.80 AUROC.^31–33^ In cSCC research, the work of Coudray et al. addresses the prediction of disease outcome from WSIs using a bag of visual words classifier, achieving AUROC=0.689 .^34^ These examples demonstrate that prediction of patient progression is indeed difficult, and that the accuracy of our model is among the best achieved so far.

Strikingly, progression risk of a patient could be predicted based on histology images alone, exceeding by far the accuracy achieved by a model trained on clinico-pathological features. Unlike clinical parameters,^7,8^ or gene expression measurements,^9,10^ which in different clinical centers might follow different standards, be done selectively for some patients only, and come with a high cost, histology is routinely performed in cSCC diagnosis. The fact that tissue slides are available for every patient and that prediction is fast and free of additional costs, considerably increases the facility and potential of our model for clinical use. Moreover, by obviating the need for data sharing, FL greatly facilitates further model training and refinement and its extension to additional centers.

Unlike prediction based on clinical parameters, which are numeric and unambiguous, prediction based on image data is not easy to interpret. Commonly, multiple instance learning models are interpreted using qualitative inspection of image regions with high attention scores.^22–24^ Here we adopt a fully quantitative and systematic approach to model interpretation in which we filter predictive patches of each patient group and statistically compare over 524 cell-based features between the two groups. Our features are based on a segmentation model specifically designed for this tumor type and capture a broad range of aspects of sample cell composition, spatial organization of the tissue, as well as nuclei morphology.^17^ We point to several noticeable differences in tumor morphology between progressing and non-progressing patients.

Interestingly, the most predictive patches of disease progression were located outside of the tumor region. In contrast, in patients without disease progression, the predictive patches were inside the tumor according to our IGs-based analysis. On the level of cellular morphology and tissue architecture, tumors from patients with disease progression exhibited a higher degree of heterogeneity. Parameters quantifying nuclear morphology showed higher variability and in these patients, cells in the tumor tissues showed a less uniform distribution. Different areas in and around the cSCC tumor, as well as features of cellular morphology may play distinct roles in the propensity for local recurrence and/or metastatic spread. Future studies in additional cohorts, ideally together with genomic and transcriptomic experiments will be instrumental to further validate our model and infer cause-and-effect relationships between morphological findings and risk of disease progression.

In summary, our study presents an explainable, federated deep learning model that reliably stratifies cSCC patients at high risk of disease progression and identifies their characteristic morphological features. The accuracy, interpretability, and federated implementation of our model hold great promise to better understand the disease and to advance the management of cSCC patients in the future.

## Methods

### Patient cohorts

For the initial training cohort, all patients with a primary cSCC diagnosed and treated by excision at the Department of Dermatology at the University Hospital Cologne (Cologne cohort) between January 2009 to May 2019 were collected. For these patients we used clinico- pathological parameters based on medical records and pathology reports and performed active follow-up regarding disease progression status. In the cohort, 96 patients experienced disease progression (metastasis and/or local recurrence), out of which histological specimens from the primary tumor for 54 patients were available. For the deep learning classification, all available progressors were used together with a random sample of primary tumors from patients without disease progression. Local recurrence or lymph-node/distant metastasis within 2 years after initial diagnosis was considered a progression event. Hematoxylin-Eosin (HE) stained slides obtained during routine work-up of surgical samples were available for 162 patients (progress n=54, non-progress n=108). From the University Hospital Bonn (Bonn cohort) patients diagnosed and treated for cSCC between March 2012 and September 2021 were included. Tumors were excised at the Department of Dermatology or the Department of Oral and Maxillo-facial Surgery and worked up histologically following standard procedures. We identified 23 primary cSCC cases with eventual disease progression (recurrence/metastasis) and randomly selected a group of primary cSCCs without disease progression. Of those, HE slides were available for 39 patients (progress n=23, non-progress n=16). For the cohort from the Department of Dermatology, Technical University Munich (TU Munich, Munich cohort) we identified patients with a primary cSCC and disease progression and assembled a random cohort of primary cSCCs without disease progression. Of those, HE slides were available for 51 patients (progress n=21, non-progress n=30). Patient inclusion and analysis was approved by the institutional review boards (Ethic vote numbers 187/16, 21- 1500, 20-1082 and 22-1330-retro).

### Analysis and classification of whole slide images

#### Datasets

Whole-slide images (WSIs) were acquired from HE slides using a NanoZoomer Slide Scanner (Hamamatsu) at 40x resolution. In total, we collected 219 WSIs of 162 patients from the University Hospital Cologne, 291 WSIs of 39 patients from the University Hospital Bonn, and 129 WSIs of 51 patients from TU Munich. We filtered out slides without any tumor tissue according to the Segmenter model described by Sancéré et al. The final dataset used for training of the federated deep learning model comprises 214 slides from 157 patients from the University Hospital Cologne, 133 slides from 35 patients from the University Hospital Bonn and 113 slides from 51 patients from TU Munich. From this dataset, 228 slides are from patients showing cSCC progression, and 232 slides are from patients showing no cSCC progression. Data splitting is done in a stratified fashion on patient level, making 65-15-20 splits for training, validation, and testing, respectively.

#### Pre-processing

Each WSI is tiled into patches of 256x256 pixels at x20 magnification. Patches without tissue are discarded, and the remaining patches are processed with an ImageNet pre-trained EfficientNet-v2-L,^35^ to compute its feature vector representations.

#### Classification

Each WSI is treated as the sequence of feature vectors corresponding to its non-empty image patches. We use the multiple instance learning classification model described by Pisula and Bozek.^36^ Following an approach similar to Lu et al.,^37^ a transformer model initialized with language-modeling pre-training weights is used for classification. We use a RoBERTa transformer encoder,^38^ and perform parameter-efficient fine-tuning by only training its normalization layers.^37,39^ To reduce compute and memory footprint, we apply multi-head attention pooling at the input to shorten the length of the patch sequence. The embedding vectors from the last layer of the transformer encoder are averaged and fed to a linear layer for the final classification.

Each WSI is classified independently during model training. During inference, in cases where there are multiple slides per patient, we evaluate the model on each one and take the prediction corresponding to the slide with the biggest activation in the positive class output neuron.

#### Model training

We train our model with a Federated Averaging strategy for 50 rounds.^12^ Adam is used as the optimizer algorithm, with a learning rate of 1.e-4, weight decay of 5.e-5, and batch size of 4. Model selection is done based on weighted validation AUROC of the three cohorts.

### Classification explanation and analysis

Beyond mere disease progression prediction with a deep network classifier, we investigate the biological features that drive our classifier’s decision. Our process is threefold: we detect relevant image regions responsible for the model’s decision; we compute handcrafted features of the cellular composition of the image regions; and we perform the data analysis itself. This approach is described in detail below.

### Input attributions

We use Integrated Gradients (IGs) to identify regions of a WSI that play a role in the classifier’s progression prediction.^16^ IGs is a deep learning explainability algorithm that attributes the prediction of a deep network to its input features. We apply IGs to our cSCC progression prediction model, to assign a positive score to image patches that contribute to the prediction of the correct class, and a negative score to patches that contribute to the prediction of the opposite outcome. By arranging the IGs attribution scores of the patches in their corresponding spatial locations in the slides, it is possible to visualize these values as heatmaps, as shown in **Fig. 3**.

### Patch description and feature engineering

We use the HoverNet nuclei segmentation model described by Sancéré et al. on the WSI image patches to identify their cell composition.^17,19^ The model detects and classifies cell nuclei into granulocytes, lymphocytes, plasma cells, stroma cells, tumor cells, and non-neoplastic epithelial cells. Once the cells in a patch have been identified, we compute a total of 524 features that summarize the patch into a single feature vector. These features include:

- Cell type populations and ratios.

- Descriptive statistics (mean, median, variance, skewness, kurtosis, minimum, maximum) of nuclei morphology, such as the mean tumor cells nuclei eccentricity, or the variance in plasma cells nuclei area. These features were computed with the ‘skimage.measur’ Python package.^40^

- Descriptive statistics of distances between cell nuclei, such as the median distance between stroma cells and tumor cells.

- Average Nearest Neighbor Ratio (ANNR) and Join Count analysis for each cell type.

The features from the last item are used to quantify the spatial arrangement of cells within a patch, and they capture two different aspects of it.

ANNR is used to quantify the observed pattern of distances between cell nuclei in a patch:

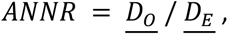

Where 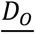 is the observed mean distance between each cell and its closest neighbor, and 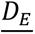 is the expected mean distance between each cell and its closest neighbor if the cells were placed randomly:

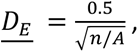

where *n* is the number of cells in a patch, and *A* is the patch area. An ANNR<1 indicates clustering (meaning, cells in the patch are closer than a random pattern of cells), and an ANNR>1 indicates a dispersed or regular pattern of cell nuclei. We compute the ANNR for each cell type in a patch.

Join Count analysis gives a measure of spatial autocorrelation: it describes how the values of a variable at neighboring spatial locations are similar to each other. In our case, the variable of interest is the cell type, where a positive spatial autocorrelation would mean that neighboring cells belong to the same type, and a negative spatial autocorrelation would mean that neighboring cells belong to different classes. Spatial autocorrelation is complementary to ANNR, it quantifies neighboring cell nuclei types disregarding how close or distanced they are. Our Join Count analysis is computed for each cell type individually, in the following way:

- A patch is partitioned into a Voronoi tessellation, using the nuclei centroids as seeds for the regions.

- The regions are binary-labeled. Given a cell type, a positive label is assigned to all the cell nuclei belonging to that class, and a negative label is assigned to the remaining regions.

- The different types of joins were then counted. Two neighboring cells make a black-black (BB) join if they both are from the positive label (i.e. the cell type being currently analyzed); a black-white (BW) join is formed between two cells of opposite labels; and a white-white (WW) join happens when two cells of the negative label neighbor each other.

This procedure is done for each cell type independently, assigning the positive label (black) to the analyzed cell type and the negative label (white) to all the other cell types. Our measure of spatial autocorrelation is given by:

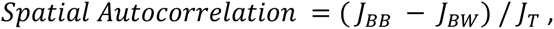

where *J_BB_*, *J_BW_*, and *J_T_* are the number of BB joins, the number of BW joins, and the total number of joins, respectively. This equation is positive when the majority of joins in a patch are BB joins, indicating a positive spatial autocorrelation, and is negative when the majority of joins are BW joins, indicating negative spatial autocorrelation.

### Data analysis

We apply IGs to all the patients in the test set, and describe their corresponding image patches as previously explained. We use in this analysis the patches coming from tumor regions detected by the Segmenter model described by Sancéré et al.,^17,41^ plus a surrounding tissue stripe of approximately 800μm of width next to the tumor border. From the totality of patches, we form two groups: A “positive group” of image patches coming from progression patients, which were detected to be explainable of this condition with IGs; and a “negative group” of patches coming from non-progression patients, which were detected to be explainable of this condition with IGs.

To enhance the predictive signal and avoid over-representing patients with bigger tumors, we take a slide’s top 10% IGs-scored patches, and limit this quantity to 200 image patches per slide. We compare values of each feature individually between the two groups of patches. We guide our analysis by focusing on features whose values differ between the two groups with an Effect Size bigger than random. We use the Common Language Effect Size (CLES),^42^ or probability of superiority, as it has no assumptions about the data distribution, and is straightforward to understand:

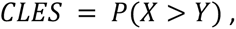

is the probability that a value sampled from group X is bigger than a value sampled from group Y. In our case, the two groups are the positive and the negative groups previously described, and we compute the CLES for each feature with brute force, by exhaustively comparing each value of one group with all the values of the same feature in the other group.

In addition to comparing the feature distributions in both groups, we tested whether the individual patches’ feature vectors were sufficient to predict the progression status of their respective patients using an XGBoost classifier.^18^ The patches under analysis were split into 80-20 train and test sets, and model selection was done with 3-fold cross-validation on the train set.

### Statistical analysis of clinico-pathological variables

Associations of clinico-pathological variables with disease progression and survival were done for all patients with available data. Association with disease progression risk was calculated using logistic regression and reported as odds ratios. Association with survival was done using the Kaplan-Meier method with log-rank test as well as Cox proportional hazard models and reported as hazard ratios with 95% confidence intervals. For multivariable analyses, variables with p<0.1 in univariate analysis were combined. Analyses were done in R statistical environment (v4.3.0).

## Data Availability

All data produced in the present study are available upon reasonable request to the authors.

## Acknowledgements

A.F. was partly funded by the Deutsche Krebshilfe through a Mildred Scheel Foundation Grant (grant number 70113307). C.L. was partly funded through the collaborative research center grant on small cell lung cancer (CRC1399, project ID 413326622) by the German Research Foundation (DFG). Both K.B. and J.I.P. were hosted by the Center for Molecular Medicine Cologne throughout this research. K.B. and J.I.P. were supported by the BMBF program Junior Group Consortia in Systems Medicine (01ZX1917B) and BMBF program for Female Junior Researchers in Artificial Intelligence (01IS20054).

