## Supplementary material for "Explainable, federated deep learning model predicts disease progression risk of cutaneous squamous cell carcinoma"

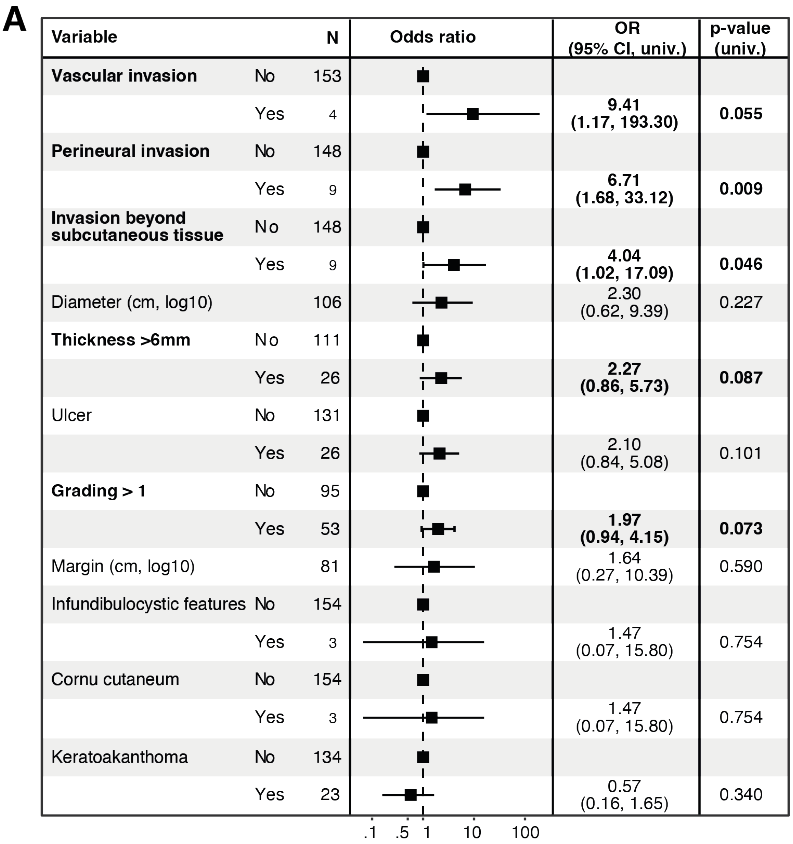


**Suppl. Figure 1:** Association of clinico-pathological parameters with cSCC progression risk calculated using logistic regression for Cologne patients with available data. Shown are Odds ratios (ORs) with 95% Confidence intervals (CIs) and univariate p-values.

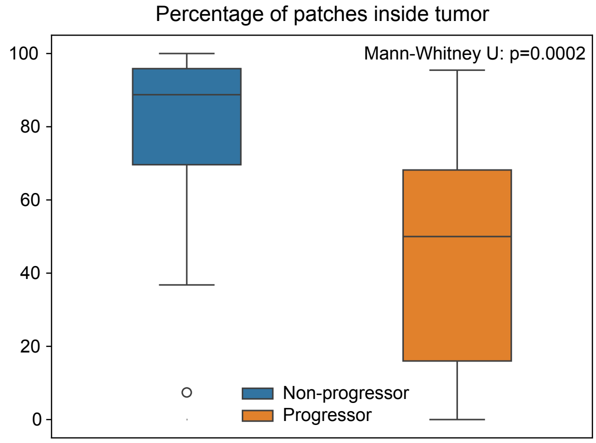


**Suppl. Figure 2:** Percentage of relevant patches (as detected by IGs) of individual patients inside the tumor regions. On average, non-progressors have more relevant patches inside the tumor compared to progressors.

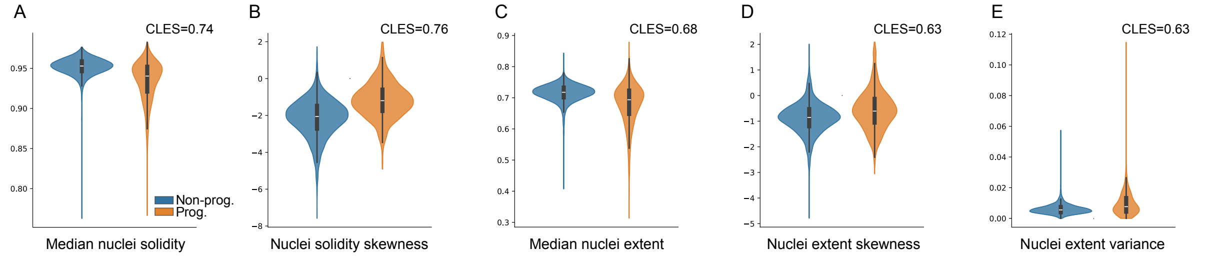


**Suppl. Figure 3:** Violin plots of 5 tumor cell nuclei morphological features. Non-progressors have larger values of morphological solidity and extent (larger median, negatively-skewed distributions, **A-D**), while morphological extent has a larger variance in tumor cells from progressors **(E)**. All features are significantly different in both groups, with p-values < 0.0001 using Mann-Whitney U test.

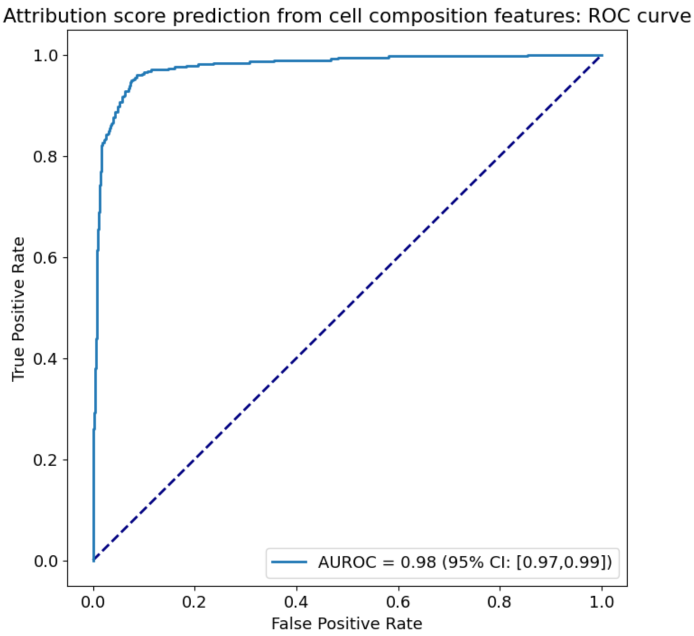


**Suppl. Figure 4:** ROC curve of the XGBoost patch-level progression status classifier using cell-based features as input.

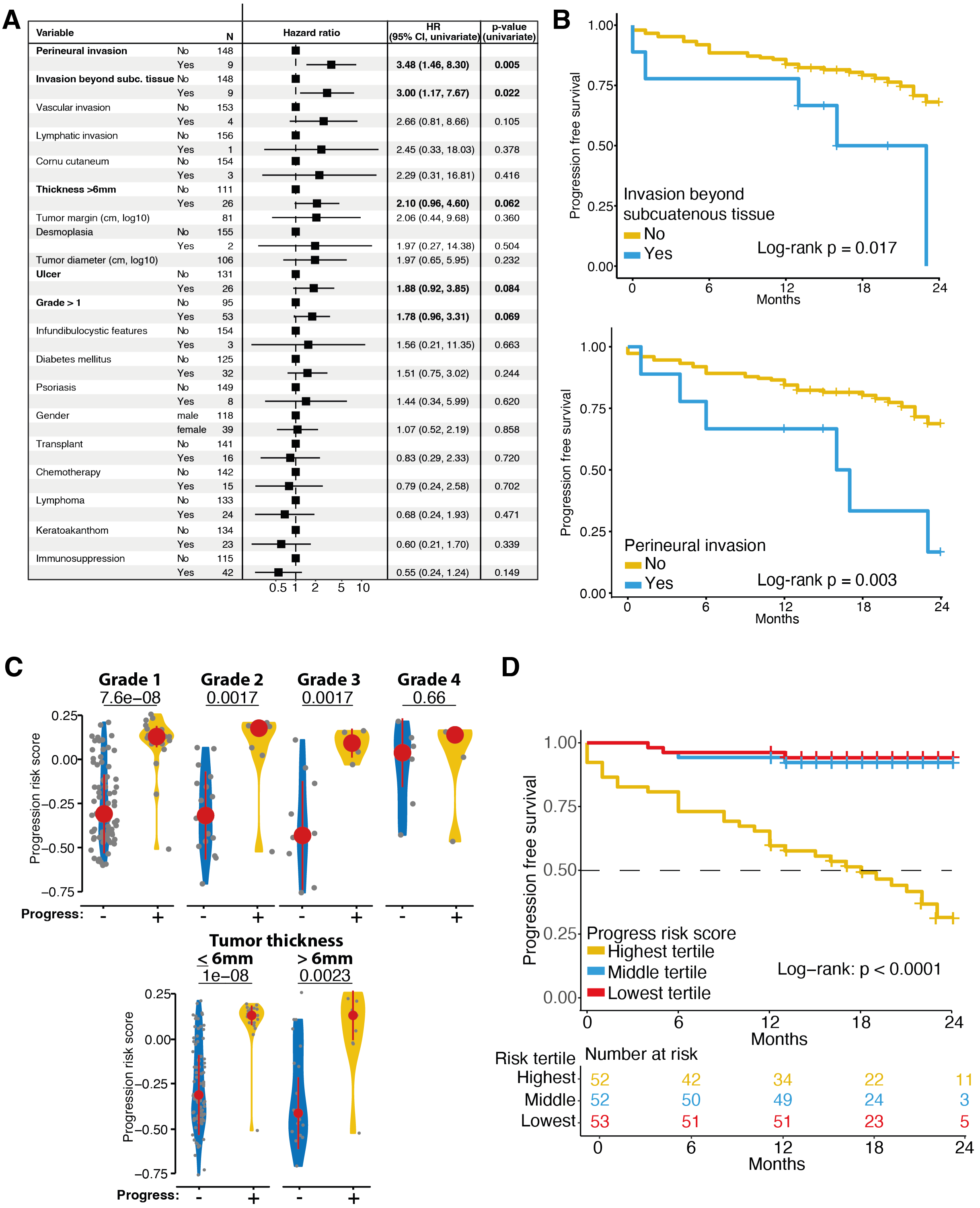


**Suppl. Figure 5: A:** Univariate association of clinico-pathological parameters derived from medical records & pathology reports with progression free survival of Cologne cSCC patients. Shown are Hazard ratio (HR) with 95% confidence interval (CI) based on Cox proportional hazard models. N indicates number of patients with available data per category. **B:** Kaplan-Meier curves for Cologne patients with or without invasion beyond subcutaneous tissue (top) or perineural invasion (bottom). **C:** Comparison of deep learning-based progression risk scores in Cologne patients with or without cSCC progression stratified by pathological grade (top) or tumor thickness >6mm (bottom). Shown are median and median absolute deviation. p-values calculated by t test. **D:** Progression free survival of patients grouped into tertiles of the deep learning-based progression risk score.

**Suppl. Table 1**

Top 100 features with the largest CLES, or probability of superiority, between the groups. To avoid displaying redundant features, pairs of features with a Pearson correlation coefficient bigger than 0.9 are grouped together, and a single feature from the group is shown. The rows are sorted in descending order of CLES for each feature type. The “Higher in” column indicates the group with larger feature values. The fraction of image patches that do not show any value for the features are shown, and features missing in more than 90% of the patches are not displayed. All the features in the table are significantly different in both groups with p<0.0001 using Mann-Whitney U test. Description of nuclei morphology features can be found in the documentation of `skimage.measure` (https://scikit-image.org/docs/stable/api/skimage.measure.html#skimage.measure.regionprops).
